# Continuing inequalities in COVID-19 mortality in England and Wales, and the changing importance of regional, over local, deprivation

**DOI:** 10.1101/2022.01.28.22270022

**Authors:** Gareth J Griffith, Gwilym Owen, David Manley, Laura D Howe, George Davey Smith

## Abstract

**Background:** Observational studies have highlighted that where individuals live is far more important for risk of dying with COVID-19, than for dying of other causes. Deprivation is commonly proposed as explaining such differences. During the period of localised restrictions in late 2020, areas with higher restrictions tended to be more deprived. We explore how this impacted the relationship between deprivation and mortality and see whether local or regional deprivation matters more for inequalities in COVID-19 mortality.

**Methods:** We use publicly available population data on deaths due to COVID-19 and all-cause mortality between March 2020 and April 2021 to investigate the scale of spatial inequalities. We use a multiscale approach to simultaneously consider three spatial scales through which processes driving inequalities may act. We go on to explore whether deprivation explains such inequalities.

**Results:** Adjusting for population age structure and number of care homes, we find highest regional inequality in October 2020, with a COVID-19 mortality rate ratio of 5.86 (95% CI 3.31 to 19.00) for the median between-region comparison. We find spatial context is most important, and spatial inequalities higher, during periods of low mortality. Almost all unexplained spatial inequality in October 2020 is removed by adjusting for deprivation. During October 2020, one standard deviation increase in regional deprivation was associated with 2.45 times higher local mortality (95% CI, 1.75 to 3.48).

**Conclusions:** Spatial inequalities are greatest in periods of lowest overall mortality, implying that as mortality declines it does not do so equally. During the prolonged period of low restrictions and low mortality in summer 2020, spatial inequalities strongly increased. Contrary to previous months, we show that the strong spatial patterning during autumn 2020 is almost entirely explained by deprivation. As overall mortality declines, policymakers must be proactive in detecting areas where this is not happening, or risk worsening already strong health inequalities.

**Key messages:**

- Spatial inequalities in local mortality are highest in periods of lower overall mortality.
- Spatial inequality in COVID-19 mortality peaked in October 2020, before decreasing strongly in November and over the winter period.
- Deprivation explains almost all inequality during October when inequality was at its highest.
- Regional deprivation was far more strongly associated with local mortality than local deprivation during September to November 2020.
- This is consistent with an overdispersed distribution of secondary infections governed by transmission heterogeneity structured by deprivation.

## Background

Throughout the pandemic many observers noted a distinct geography to SARS-CoV-2 infection and subsequent COVID-19 disease outcomes in England, with substantially worse outcomes observed in more deprived areas. Previously, we used multilevel modelling to measure how geographical inequalities in Covid-19 mortality changed in England over the first five months of the pandemic from March until July 2020 [1]. We partitioned unexplained inequalities at three different geographical scales from a local neighbourhood scale up to a wider region.

This initial paper had four main findings: firstly, we found substantially greater geographical inequalities in COVID-19 related mortality than in non-COVID mortality. Secondly, while there was some variation depending on the geographical scale, we broadly found that inequalities increased over the study period and were greater when absolute mortality was lower. Thirdly, we presented evidence that area level deprivation was associated with higher COVID-19 related mortality, however after adjusting for local deprivation, inequalities in COVID mortality remained largely the same. Finally, we found substantial inequalities between geographical units at all three scales in our analysis.

In this paper we extend and refine our early analysis using newly available data reporting deaths due to COVID, rather than deaths involving COVID, using small areas (Middle Layer Super Output Areas, MSOAs) up until April 2021 [4]. Daily deaths peaked at 1270 deaths in England and Wales on January the 19^th^ before declining rapidly during spring 2021. At the end of the study period, in April 2021, there were around 30 daily deaths within 28 days of a positive COVID-19 test. However, understanding how inequalities within this aggregated narrative played out over the period remains very relevant. Understanding past spatial patterns helps contextualise current spatial inequalities [5,6] and helps policymakers ensure inequality from COVID-19 mortality is kept to a minimum.

The context of the COVID-19 pandemic changed dramatically over the period from July 2020 to April 2021. In Summer 2020, the end date of the previous data release, case rates were low in England and many restrictions had been eased, although some local restrictions were implemented where case rates were rising or remained high [7]. At the beginning of Autumn 2020, however, cases started to rise rapidly. At this time cases were unevenly distributed across the country with a notable North-South divide. As a response the UK government introduced a tiered system in late October [8] in which local authorities were allocated to one of three tiers with restrictions of increasing stringency. The tier to which a local authority was allocated was based on a range of factors including case numbers and pressure on the local health service [9]. The whole of England entered a 27-day lockdown from the 5th November before a return to an updated three-tier system at the beginning of December. A further rapid rise in cases, hospital admissions and deaths preceded the whole country being placed in a national lockdown on 6th January 2021 [8]. After the January 19^th^ mortality peak case numbers and deaths declined, and starting from March 2021 restrictions were gradually eased over the rest of the study period. Figure 1 shows the evolution of combined English and Welsh mortality over the period, highlighting the period of tiered restrictions, and the 27-day national lockdown which was imposed during this period.

**Figure 1.**
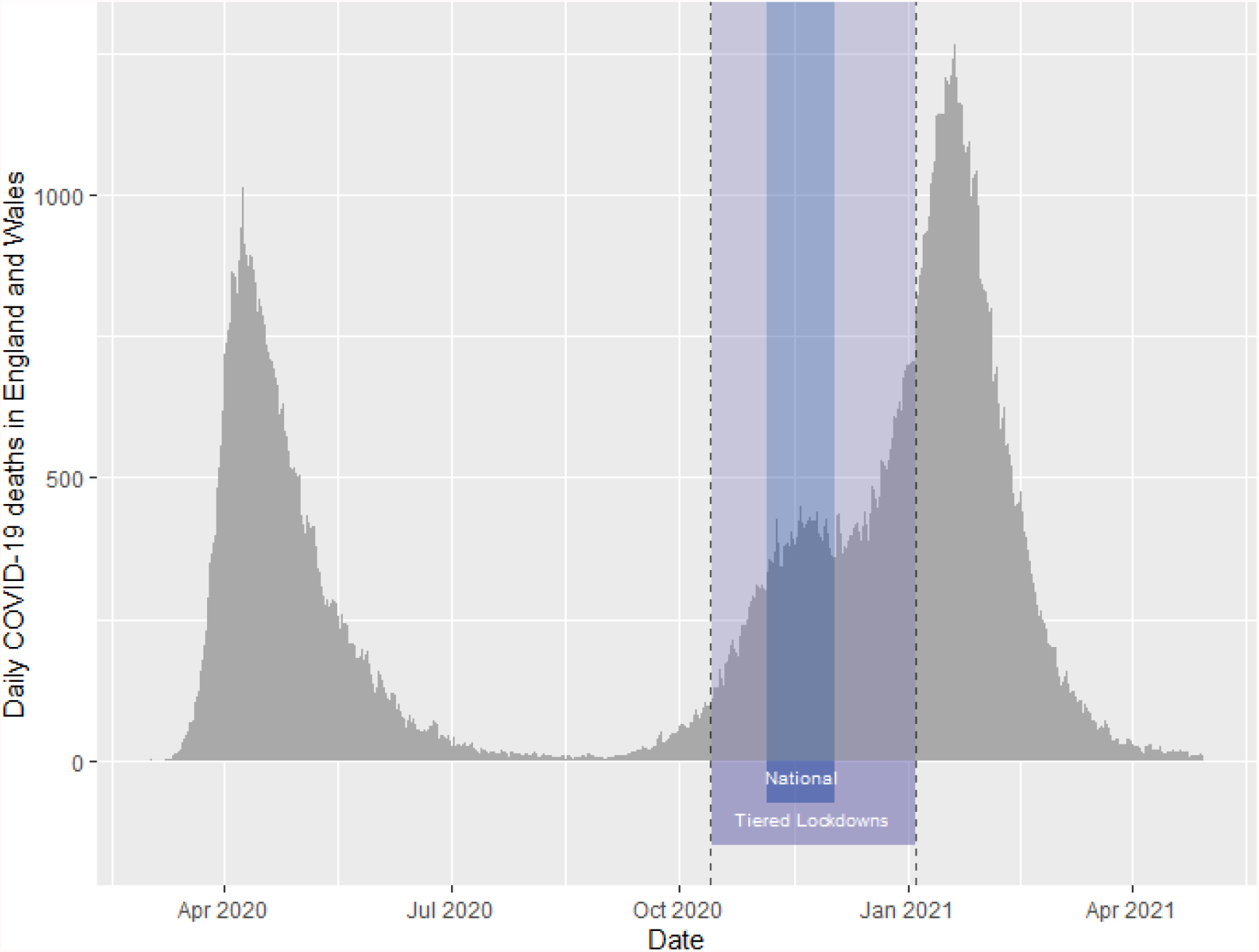
Number of deaths recorded with COVID-19 as cause of death per day in England and Wales, over the study period. Purple highlight indicates the period of locally sensitive tiered restrictions. Blue highlight indicates 27 days of national lockdown from 5^th^ November to 2^nd^ December 2020.

Research has consistently shown that neighbourhood deprivation is associated with higher COVID-19 case rates [10–12], and recent work suggests this was more true between August and October 2020, than previously. However, this association with deprivation was altered after the first period of localised tiered restrictions, with the most deprived areas reporting lowest COVID-19 case rates in November and December [2]. As yet, there is a lack of work investigating the extent to which this reversal was reflected in mortality, and furthermore how this change was structured geographically. For instance, what was the relative importance of local versus regional deprivation on local COVID-19 mortality.

In the light of newly available data, and the changing COVID context over the study period, here we present new analyses, focusing particularly on three principal research questions:

1. Do geographical inequalities in COVID-19 mortality continue to exceed geographical inequalities in non COVID-19 mortality?
2. Is higher overall COVID-19 mortality still associated with lower spatial inequality in COVID-19 mortality?
3. Is there a discernible impact on the relationship between deprivation and local mortality during the period before and after local tiered lockdowns were implemented?

## Methods

### Data

All data used here are publicly available from the Office for National Statistics (ONS), Environmental Systems Research Institute, or derived data sets from existing published research. Data on care homes, age structure, and geographical identifiers are consistent with our work on deaths involving COVID-19 [1], and we provide open access links here. We give information below on data which differ from the initial manuscript, describing the ONS reporting change in mortality counts, and an update of the UK equivalised Index of Multiple Deprivation (UKIMD).

### Outcome

Mortality data are counts of age-standardised MSOA-level mortality from COVID-19 and non-COVID-19 sources, with data now available from March 2020 to April 2021. In November 2020, the ONS changed COVID-19 mortality reporting. Mortality data now represent “deaths where COVID-19 was the underlying cause of death”[4], rather than “deaths where COVID-19 was recorded on the death certificate” [13]. For our purposes this represents an improvement, as we are interested in clustering of deaths due to COVID-19, not in clustering of deaths involving COVID-19. High numbers of deaths involving COVID-19 may indicate high COVID-19 prevalence amongst those who died, rather than that such COVID-19 prevalence caused death. We estimate our models over the entire period using this improved classification. As such, where March to July 2020 estimates differ from previous work, this may indicate structural differences in initial death misclassification.

We repeat all analyses for “non-COVID mortality”, inferred from deaths where COVID-19 is not listed as a cause of death. We treat non-COVID mortality as a comparator group to explore whether any spatial structure detected in our results is unique to COVID-19 mortality or capturing spatial variation in all-cause mortality. Whilst this cannot be considered a true negative control [14], as it is not independent of our outcome of interest due to competing risks, it can help us contextualise the spatial structure of COVID mortality.

### Structural Levels

Investigating higher level spatial inequalities requires specifying plausible geographical scales at which mortality inequalities might be expressed. We operationalise local neighbourhoods using Middle Layer Super Output Areas (MSOAs, n=7201). MSOAs are nested within ONS Travel to Work Areas (TTWAs, n=173) which are constructed to represent local labour market areas, and taken to characterise commuter patterns [15]. TTWAs are in turn nested within Governmental Office Regions (GORs, n=10), these are taken to pattern the macro-scale health exposures suggested to predict COVID-19 mortality throughout the pandemic [16,17].

### Covariates

COVID-19 mortality, and local geography, are strongly spatially confounded and as such we adjust for two principle confounders. We adjust for local age structure, due to the known very strong relationship between individual age and mortality risk [18], otherwise we would likely pick up spatial structure which reflects the location of elderly individuals. Similarly, we adjust for the number of care homes, as these were (at least in the UK early pandemic context) the strongest area level predictor of mortality [3,16].

We use an updated version of the equivalised UKIMD using the method proposed by Abel et al [19] which was recently made publicly available using indicators from 2020 [20]. UKIMD coefficients are partitioned into between- and within-area measures, such that “regional UKIMD” is the regional average, “TTWA UKIMD” is the TTWA average UKIMD minus the regional average, and “MSOA UKIMD” is MSOA UKIMD minus the TTWA average. This means that, for instance, MSOA UKIMD coefficients are interpreted as the effect of a given MSOA deprivation exposure over and above that experienced due to residing in the TTWA and region that MSOA is in.

### Statistical Analyses

We specify a multilevel Poisson model to analyse MSOA mortality counts, with a geographically invariant, month-specific log-offset. This offset represents our theoretical statistical landscape in which the processes governing local mortality in each month are assumed to be spatially uniform. This measure is derived from the product of the monthly England and Wales age-standardised mortality rate and the MSOA population. Any deviation away from this theoretical distribution suggests spatial inequality in mortality. Statistically, this implies that any overdispersion (or clustering) in random coefficients under this specification allow us to infer spatial inequality in mortality, over and above that which would be predicted simply by larger population and stochastic Poisson variation [21].

The empirical summaries of the random components of our model are given by the Median Rate Ratio (MRR), which presents the median relative change in mortality rate between randomly sampled pairs of lower level units within the same higher level context [22,23]. This is our measure of inequality, with higher MRRs indicating greater levels of inequality. We calculate month-specific MRRs for each geographical level, for both COVID-19 and non-COVID -19 mortality. These are complemented by the Variance Partitioning Coefficient (VPC) which presents the proportion of supra-individual unexplained heterogeneity explained by a given level [24].

We are also interested in how the relationship between deprivation and COVID-19 mortality changed over the study period. Moreover, we are interested in whether it is more important whether a neighbourhood is deprived compared to local neighbourhoods, or whether it is the fact that the neighbourhood sits within a deprived region that drives observed associations. As such, we present regression coefficients for the association between deprivation and local mortality, structured by between- and within-area deprivation. This formulation allows us to tease apart the relative contribution of local versus regional deprivation [25].

We use Markov chain Monte-Carlo (MCMC) estimation, with a discarded burn-in of 50,000 iterations, and a subsequent monitoring chain of 500,000 iterations. Credible intervals for all estimated quantities are the 2.5th and 97.5th percentiles of the posterior parameter distributions. All models were run in MLwiN V3.09, with output extracted and processed in R. Code for post-processing is all publicly available at: https://github.com/Zimbabwelsh/covid_mort_ineq_apr21

Results are presented for both COVID-19 related and non-COVID-19 related mortality for all models. All models include fixed effect adjustment for MSOA age structure and number of care homes, with further adjustment for UKIMD in latter results.

## Results

Our first question addresses whether the magnitude of spatial inequalities in COVID-19 mortality continues to exceed that of non-COVID-19 mortality.

Figure 2 presents the development of month-specific COVID-19 mortality MRRs across the three spatial scales over the study period. The MSOA MRR for COVID-19 mortality peaks in September 2020, at 2.90 (95% CI 2.58 to 3.28), which implies that within TTWA, between-MSOA rate ratios typically imply an almost threefold relative increase in COVID-19 mortality. The TTWA peak is earlier, in August, at 2.99 (95% CI 2.46 to 3.82). The largest COVID-19 mortality MRR, however, is seen in October between regions, with an MRR of 5.86 (95% CI 3.31 to 19.00), implying that in October the median comparison between regions implied an almost 6-fold relative increase in mortality. Although these peaks are expressed in different months at different spatial scales, there is still a marked decline in inequality across all scales during subsequent months until early 2021 prior to a slight uptick in inequality in April 2021.

**Figure 2.**
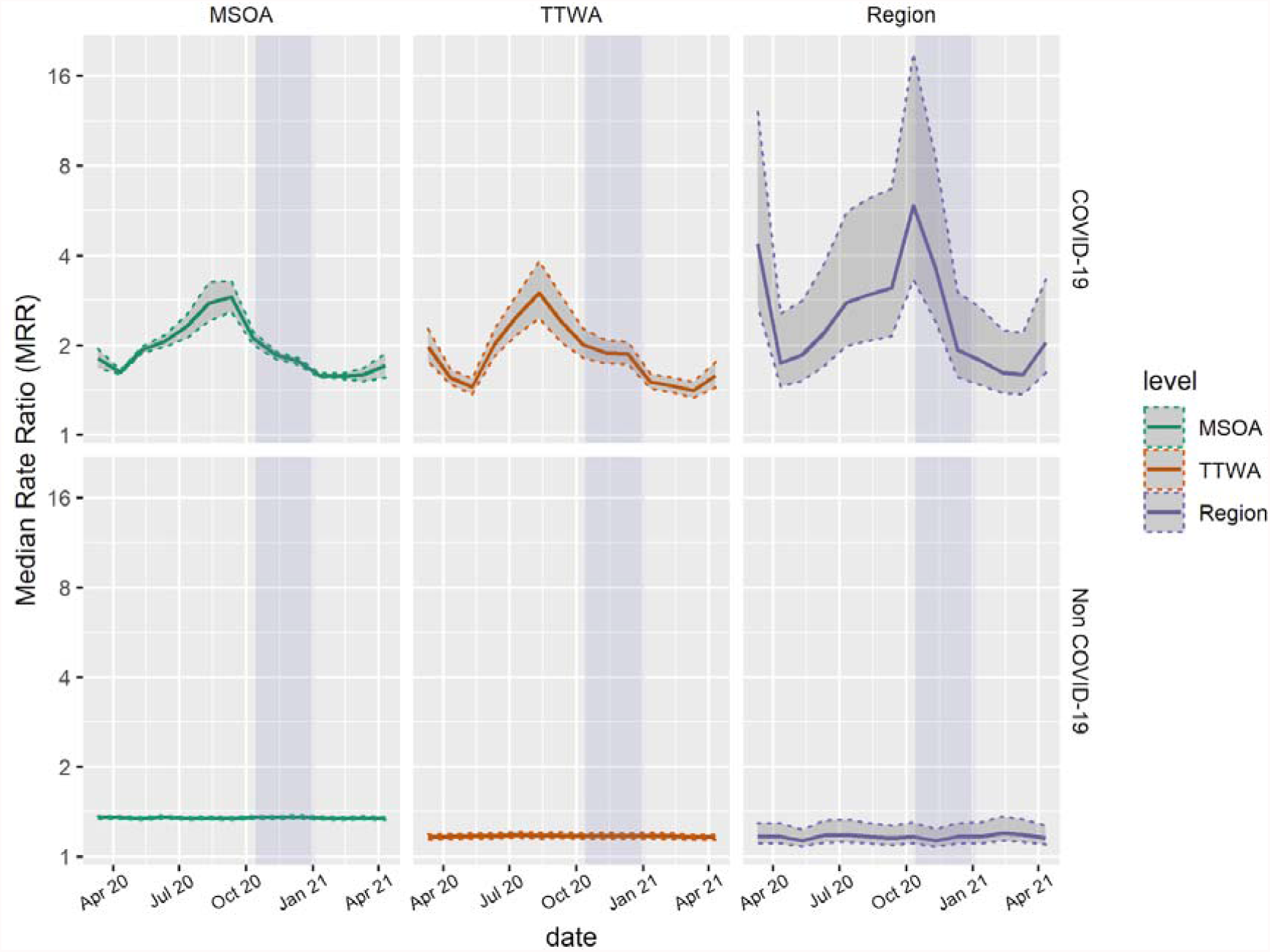
Estimates of median monthly COVID-19 and non-COVID-19 mortality rate ratios across three administrative scales from March 2020 to April 2021. Dotted intervals indicate 2.5th and 97.5th percentile credible intervals of posterior parameter distributions. Purple highlight indicates the period of locally defined tiered restrictions. MSOA, Middle-Layer Super Output Area; TTWA, Travel to Work Area.

The contrast with spatial inequalities in non-COVID-19 related mortality in Figure 2 is striking. There is very little spatial structure to non-COVID-19 mortality after adjusting for age structure and number of care homes. For all months, absolute non-COVID-19 mortality is higher than COVID-19 mortality (supplementary table 1), but the distribution of that mortality is far more spatially random, with the largest monthly mortality MRR for December 2020 at 1.36 (95% CI, 1.34 to 1.37).

We find strong evidence that during summer and autumn 2020, COVID-19 mortality inequality was at its greatest, at all spatial scales. The relative importance of each spatial scale within each month is presented in Figure 3. As expected, MSOA, as the lowest spatial scale considered explains the largest proportion of variance in all months. However, leading up to the introduction of tiered restrictions - where we see the greatest absolute inequalities, the nature of these inequalities became more strongly regional, with region explaining 37% of the total supra-individual heterogeneity in COVID-19 mortality in October 2020. This regional inequality spikes in September and October, before declining sharply again during the period of tiered restrictions in November. Again, strikingly, local non-COVID-19 mortality is explained almost entirely by the MSOA itself, the broader spatial context of the MSOA explains close to nothing.

**Figure 3.**
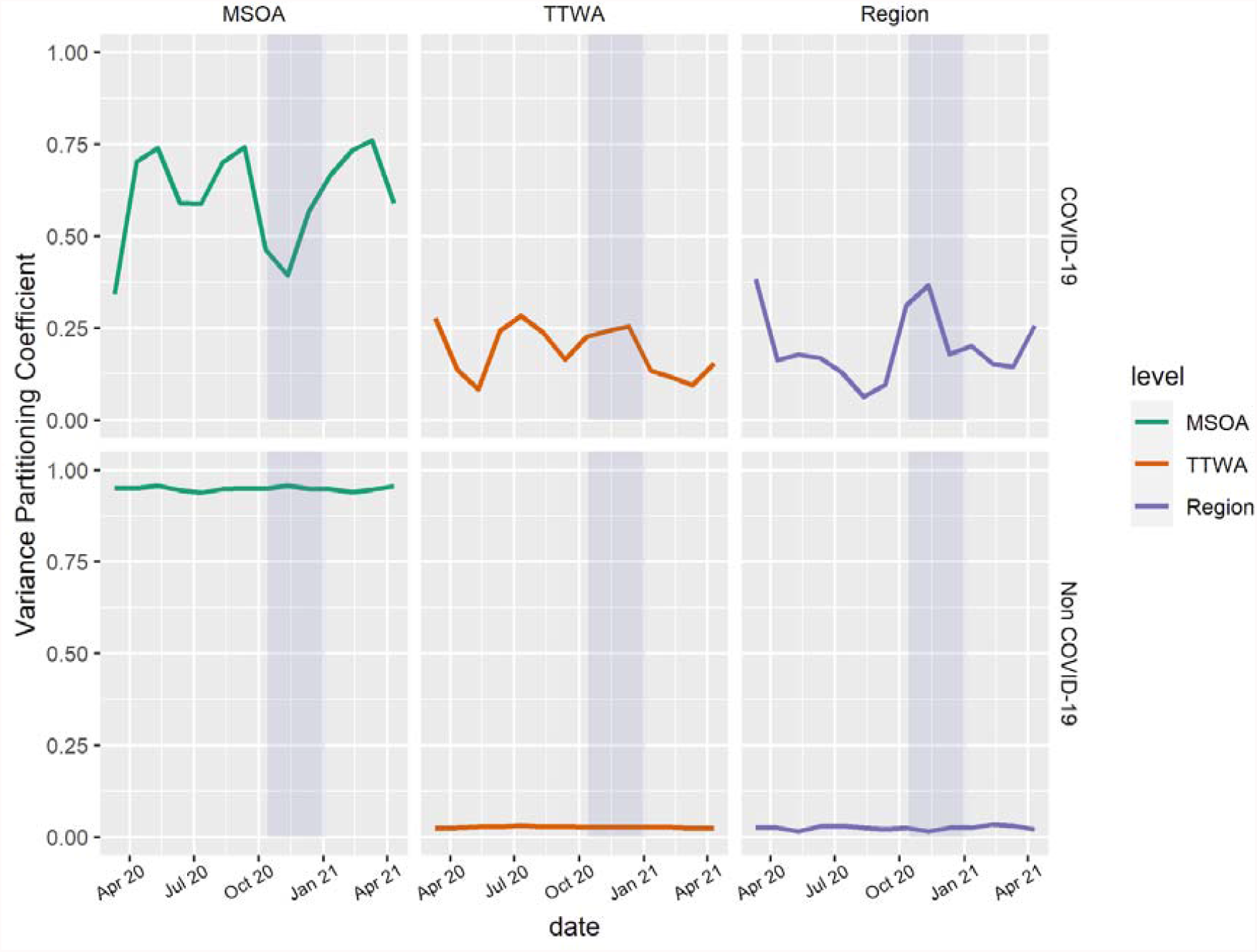
Estimates of monthly COVID-19 and non-COVID-19 VPCs across three administrative scales from March 2020 to April 2021. Purple highlight indicates the period of locally defined tiered restrictions. MSOA, Middle-Layer Super Output Area; TTWA, Travel to Work Area.

We interrogate the nature of this regional inequality change more explicitly by inspecting modelled regional residuals. Figure 4 gives the evolution of region-level mortality rate ratios over time. We can see that in the period leading into the tiered restrictions, COVID mortality was increasing strongly in the northern regions, particularly the Northeast, Northwest and Yorkshire and Humber regions. Similarly, over this period, we see strongly negative residuals for the Southwest and London. This difference peaks in October, giving rise to the MRR seen in Figure 1, where the median comparison between these rates implies a 5-fold increase in mortality. During tiered restrictions, we see rapidly declining mortality in northern regions and a strong increase in rates in London and the Southeast.

**Figure 4.**
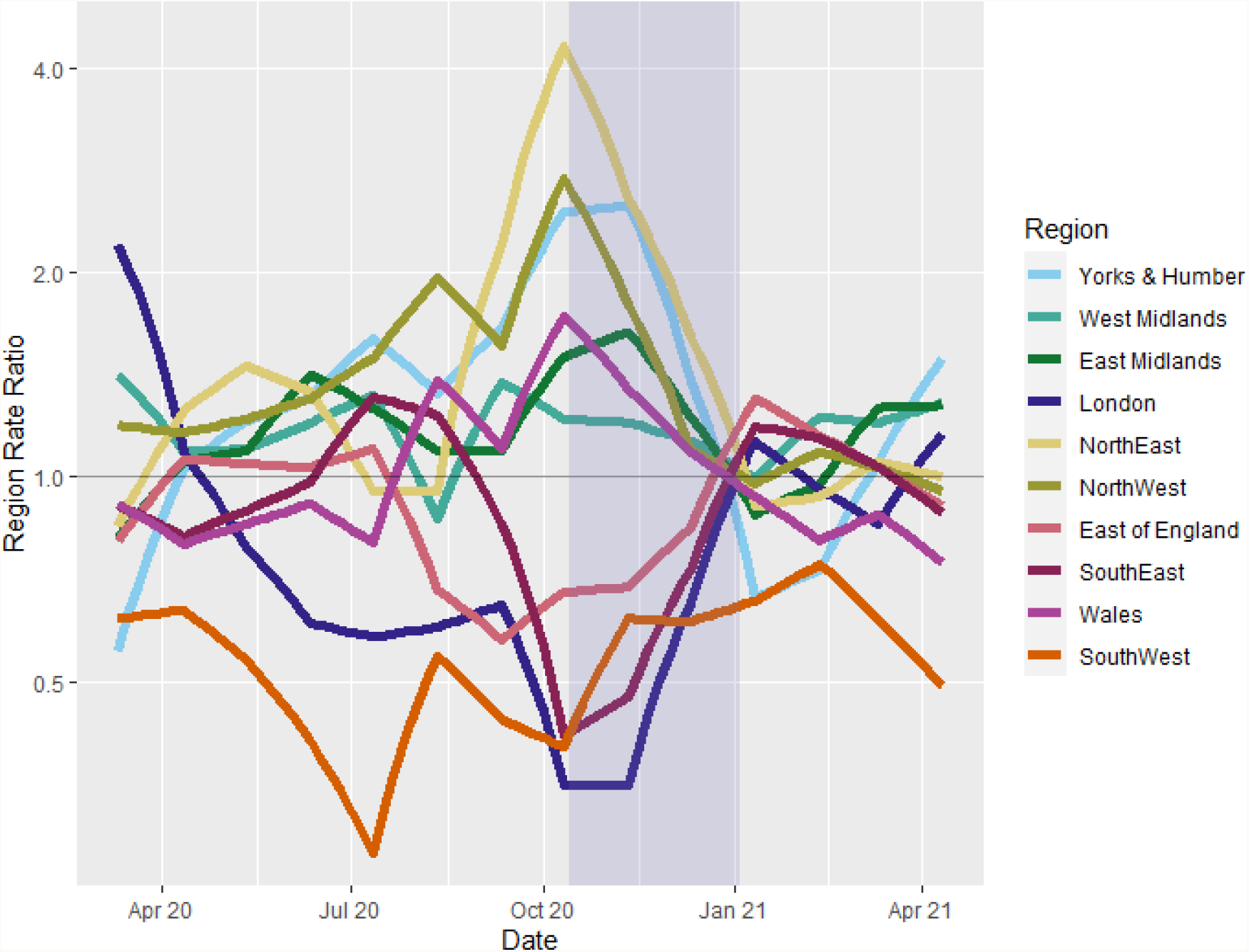
Monthly, regional COVID-19 mortality rate ratios, indicating regional mortality rate relative to precision weighted monthly population average. Model adjusted for local age structure and number of care homes. Purple highlight indicates the period of locally defined tiered restrictions.

Our second research question asks whether periods of high national mortality tend to associated with lower spatial inequalities, and vice versa. Figure 5 presents a comparison of the change in COVID-19 mortality inequality and COVID-19 mortality over the study period. We can see that mortality peaks in April 2020 and January 2021 are associated with the periods of lower spatial inequalities. Similarly, the period of greatest inequality in October 2020, is concurrent with low absolute mortality relative to the January 2021 peak. This relationship seems to hold true as mortality begins to increase in winter 2020.

**Figure 5.**
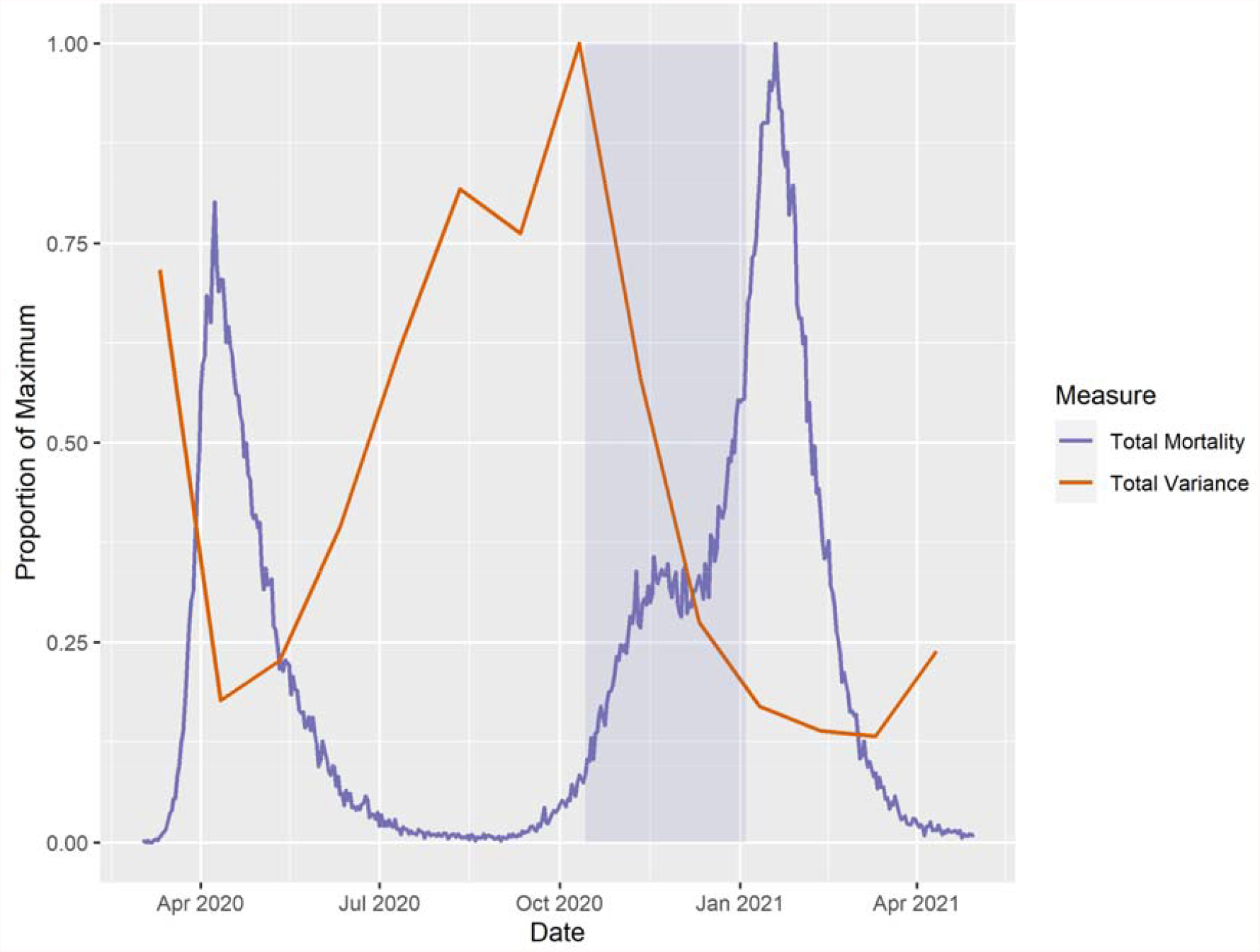
Evolution of COVID-19 mortality inequality, summed across variance components, as a proportion of maximum spatial inequality (in October 2020), and mortality due to COVID-19 in England and Wales as a proportion of January 19^th^ 2021 peak. Purple highlight indicates the period of locally defined tiered restrictions.

Having established this peak in spatial inequalities, we are still interested in the degree to which deprivation explains both COVID-19 mortality and COVID-19 mortality inequality during the study period. Figure 6 presents the MRRs from the model where we adjust for the recently derived UKIMD metric. The peak in regional inequality in October, prior to the introduction of tiered restrictions, is strongly attenuated after adjusting for local deprivation (MRR = 1.97, 95%CI 1.56 to 3.27). This differs strongly from previous research, which suggested that whilst being associated with increased local mortality, local deprivation explained very little of the unexplained spatial inequality [1]. This result is consistent with the updated VPC plot where MSOA accounts for 98.7% of unexplained spatial variation in the month of October when adjusting for UKIMD (Supplementary Figure 1). Similarly, regional residuals having adjusted for deprivation imply far less inequality (Supplementary Figure 2).

**Figure 6.**
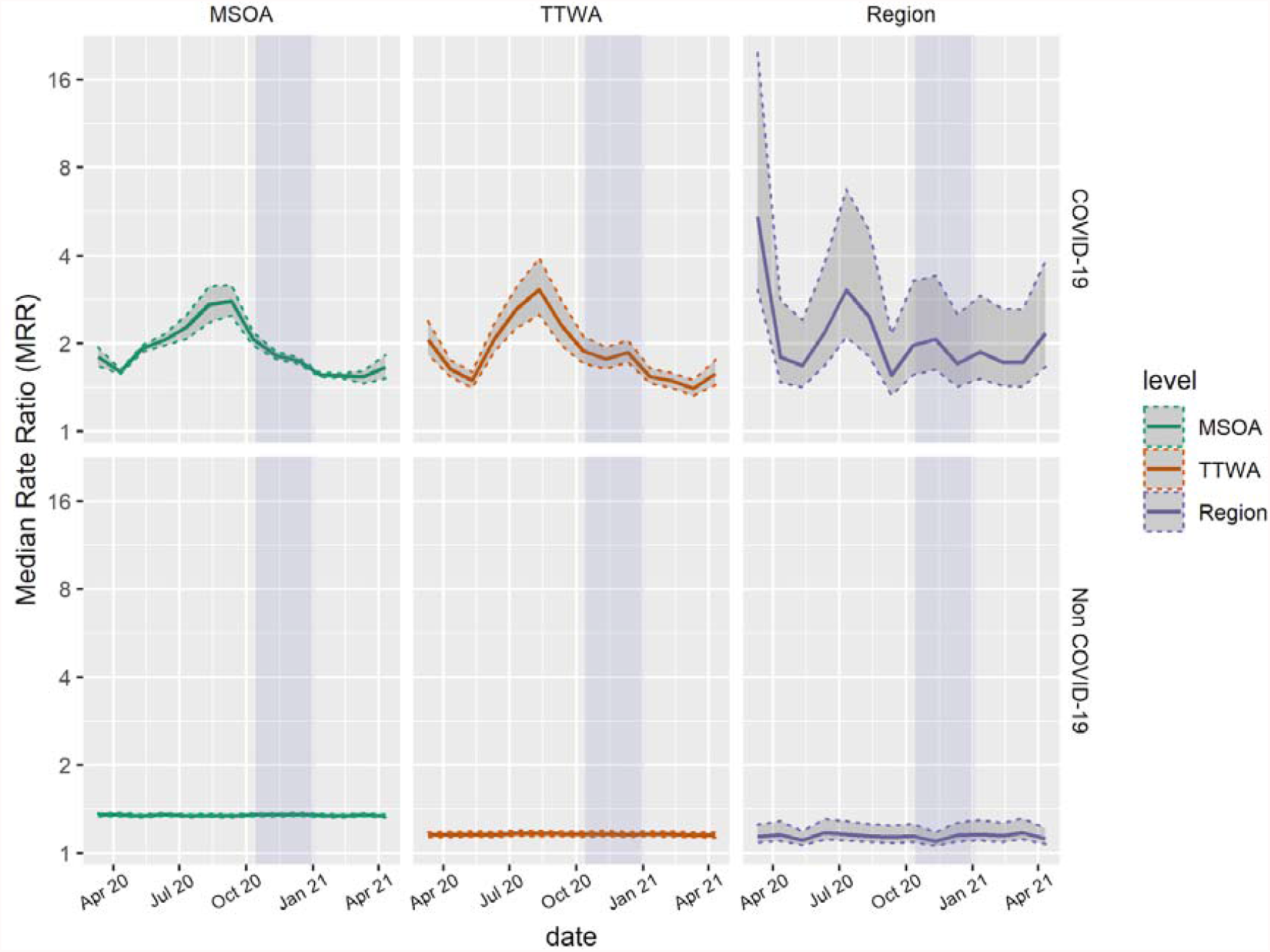
Estimates of median monthly COVID-19 and non-COVID-19 mortality rate ratios across three administrative scales from March 2020 to April 2021 after adjustment for UKIMD. Dotted intervals indicate 2.5th and 97.5th percentile credible intervals of posterior parameter distributions. Purple highlight indicates the period of locally defined tiered restrictions. MSOA, Middle-Layer Super Output Area; TTWA, Travel to Work Area. See supplementary Figure 1 for accompanying VPCs.

To further expand upon this deprivation relationship, we present the regression coefficients for UKIMD on mortality at all levels. Figure 7 presents this coefficient at three spatial scales. The association between deprivation and COVID-19 mortality increased between July and October 2021. This was true for all three geographic scales, however the increase was much more dramatic at the region level. A standard deviation increase in regional UKIMD was associated with 1.26 times higher COVID-19 mortality in July (95% CI, 0.66 to 2.25), which by October had reached 2.45 (95% CI, 1.75 to 3.48). The association between region level deprivation and Covid-19 mortality, then declines just as dramatically in the following months and by January 2021 the association between region level deprivation and mortality becomes negative. Deprivation remains associated with increased COVID-19 mortality at MSOA level at all time points. The relationship between deprivation and non-COVID-19 mortality also remains consistent throughout the study period.

**Figure 7.**
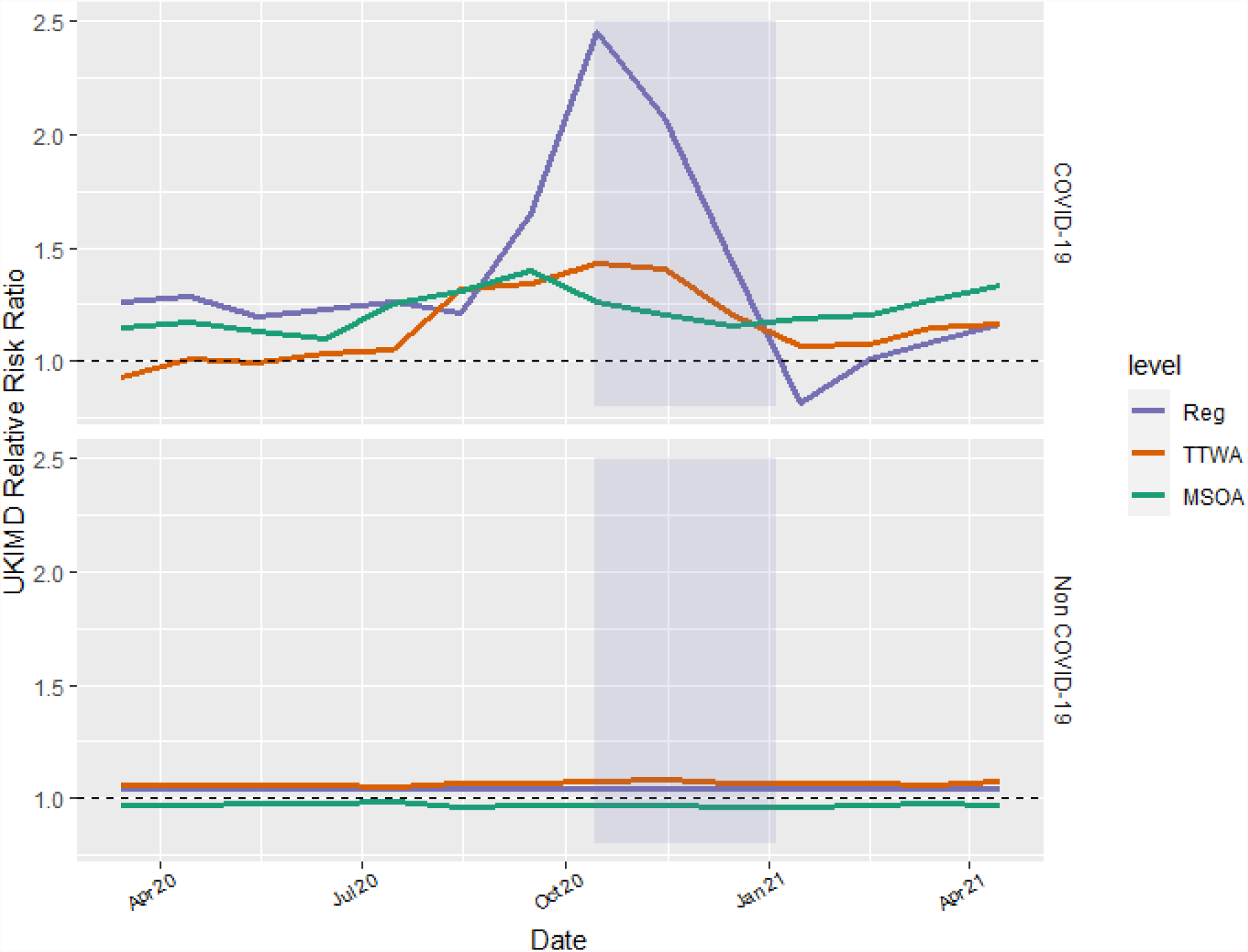
Monthly Risk Ratios for standardised UKIMD scores, for regional average, TTWA minus regional average (TTWA) and MSOA minus TTWA average (MSOA) UKIMD values. Purple highlight indicates the period of locally defined tiered restrictions. MSOA, Middle-Layer Super Output Area; TTWA, Travel to Work Area.

## Discussion

Our principal finding is that geography continues to matter a great deal for COVID-19 mortality and matters more than it does for non-COVID-19 mortality. Whilst the importance of specific spatial contexts varied, there were consistently far greater differences between places for COVID-19 mortality than for mortality from other causes.

A key finding of earlier work was that spatial inequality appeared to be broadly inversely related to countrywide COVID-19 mortality [1]. When COVID-19 mortality was at its initial peak in April 2020, mortality was high everywhere, with comparatively small differences between areas of the country. It was in the following months when mortality overall declined, that we saw spatial inequalities emerge. In this period there were very few deaths in some areas while mortality remained persistently high in others. We wanted to probe whether this had remained the case over the extended study period, and under the updated COVID-19 mortality definition from the ONS. Our findings suggest that as the pandemic progresses this association still holds, with inequalities structuring mortality burden more strongly in periods of low mortality. The COVID-19 mortality peaks in Figure 1 coincide with a reduction in inequality, seen in Figures 2 and 5. Again, we caution that whilst clearly reduction in deaths is desirable, we should remain vigilant to contextual differences in such reductions, as deaths may continue to concentrate in certain (likely disadvantaged) areas, as nationwide mortality decreases.

The most striking change in geographical inequality occurs in the period around October 2020, when regional inequality peaks, before sharply falling. This fall occurs during the period of geographically targeted tiered restrictions. We are not testing whether the decline in inequality was a direct consequence of tiered restrictions but note that it is concurrent with a small reduction in deaths in regions under the greatest restrictions. Similarly, research suggests that tiered restrictions were effective at reducing inequality in local case numbers [9]. Figure 4 clearly shows the convergence of regional mortality residuals during this period, but this is not solely driven by the mortality reduction in northern regions, where local restrictions were tighter, but also due to the increases in mortality in the South East, following the emergence of the SARS-CoV-2 lineage B.1.1.7 [26].

Our study has several limitations. We only measure the impact of geography in a strictly hierarchical sense, where knowing the higher-level context of each lower-level unit. Our model cannot consider spatial contiguity or proximity or spatial networks. Similarly, if we have mis-specified structural levels at which spatial inequalities in COVID mortality are experienced, we might expect higher-level MRRs to underestimate true inequalities. We also cannot infer about the importance of spatial context relative to within-MSOA, between-individual differences as individual-level data are not accessible [27].

There is good reason to suspect an association between deprivation and COVID-19 outcomes, for instance, labour markets in more deprived areas lend themselves less readily to working from home [28]. Here, we find strong evidence that material deprivation is associated with COVID-19 mortality, and that this association strengthened markedly between August and October 2020, supporting previous work demonstrating this with infections [2]. However, here we go further and decompose deprivation into deprivation relative to neighbouring areas, centred on the broader regional average, and find that regional deprivation is actually more informative for MSOA mortality than the relative deprivation of the MSOA itself. Moreover, we find results consistent with Morrissey et al. [2] that this relationship subsequently reverses, though reversal is delayed for mortality compared with cases. Due to our multilevel decomposition, we can also offer more insight into the relationship, and demonstrate that deprived TTWAs within regions, and deprived MSOAs within TTWAs have higher risk of COVID-19 mortality throughout the study period. Had we solely considered MSOA-level outcomes, we would have concluded that more deprived MSOAs truly had lower COVID mortality in early 2021, rather than contextualising this against a broader regional trend of increasing mortality in the southeast with lineage B.1.1.7.

Despite a strong association with mortality, deprivation does not explain geographical inequalities in COVID-19 mortality for the earlier half of the study period. However, we find that the increase in regional inequality prior to the period of tiered restrictions is almost entirely explained by deprivation. Beyond this peak, however, our results are similar to that from previous work that, for most of the study period, deprivation did not explain all or even most of the geographical inequality in COVID-19 mortality.

Taken together, the fact that regional deprivation is more strongly associated with MSOA COVID-19 mortality than the relative deprivation of the MSOA itself within the region, and that deprivation seems to predominantly explain regional inequalities at the end of the period of weakest pandemic restrictions (October 2020) seems consistent with an understanding of SARS-CoV-2 infection, COVID-19 disease and subsequent mortality as governed by time-dependent heterogeneity in transmission [29].

It was documented early on in the global spread of COVID-19, that secondary transmissions are heavily overdispersed, i.e. that most cases result in zero subsequent infections, but a small number of cases result in very high numbers of subsequent infections [30,31]. The differences between numbers of secondary infections are seemingly (at least epistemologically) stochastic; we can’t explain what causes shifts in the distribution of transmission proportion in some contexts and not others [32]. Whilst it is intuitive that deprivation predicts greater likelihood of exposure to COVID-19 disease, it seems plausible that the population averaged effect of this is possibly small relative to the inherent heterogeneity within COVID-19 transmission. This is consistent with our results where deprivation, while clearly important, only explains a small amount of total inequality for most of the study period.

This does not immediately explain the large increase in regional inequality during summer 2020, however, we find that this increase is associated with regional deprivation. In periods where case rates are low, we would expect to find larger spatial inequalities between small areas as a result of small shifts in the distribution of the transmission proportion. Moreover, we would expect that these were not intimately tied to MSOA characteristics. However, if transmission heterogeneity were structured with respect to areal deprivation, the degree of spatial aggregation required to detect such a deprivation effect would increase as a function of the inherent stochasticity. This suggests that during the summer as mortality declined, it did so in a fashion which became more strongly clustered, and that this clustering may be due to transmission heterogeneity being patterned by areal deprivation, which manifested as regional inequalities in COVID-19 mortality.

In conclusion, our study demonstrates high levels of inequality in COVID-19 mortality between places in England. Of concern looking forward, is the finding that inequalities have increased when deaths are lower. As COVID-19 likely transitions to endemicity, supplemented by high levels of vaccination in the population and far lower case fatality rates, our research highlights the imperative for policymakers to focus resources on limiting geographical inequalities. Lower rates of vaccination in deprived areas [33,34] combined with the heterogeneous nature of secondary infections [29], underpins a risk that COVID-19 worsens the already large geographical health inequalities in mortality.

## Data Availability

All data produced are available online:
ONS Deaths due to COVID-19 mortality data are available at: https://www.ons.gov.uk/peoplepopulationandcommunity/birthsdeathsandmarriages/deaths/datasets/deathsduetocovid19bylocalareaanddeprivation
ONS Deaths involving COVID-19 mortality data are available at:
https://www.ons.gov.uk/peoplepopulationandcommunity/birthsdeathsandmarriages/deaths/datasets/deathsinvolvingcovid19bylocalareaanddeprivation
ONS Administrative geographical identifiers are available at:
https://geoportal.statistics.gov.uk
ONS population health estimates are available at: https://www.ons.gov.uk/peoplepopulationandcommunity/populationandmigration/populationestimates/datasets/middlesuperoutputareamidyearpopulationestimates.
ESRI care home estimates are available at: https://covid19.esriuk.com/datasets/e4ffa672880a4facaab717dea3cdc404_0
Equivalised UK IMD estimates are available at:
https://github.com/mysociety/composite_uk_imd
All code to reproduce analyses is available at:
www.github.com/zimbabwelsh/covid_mort_spatseg

---

**Supplementary Figure 1.**
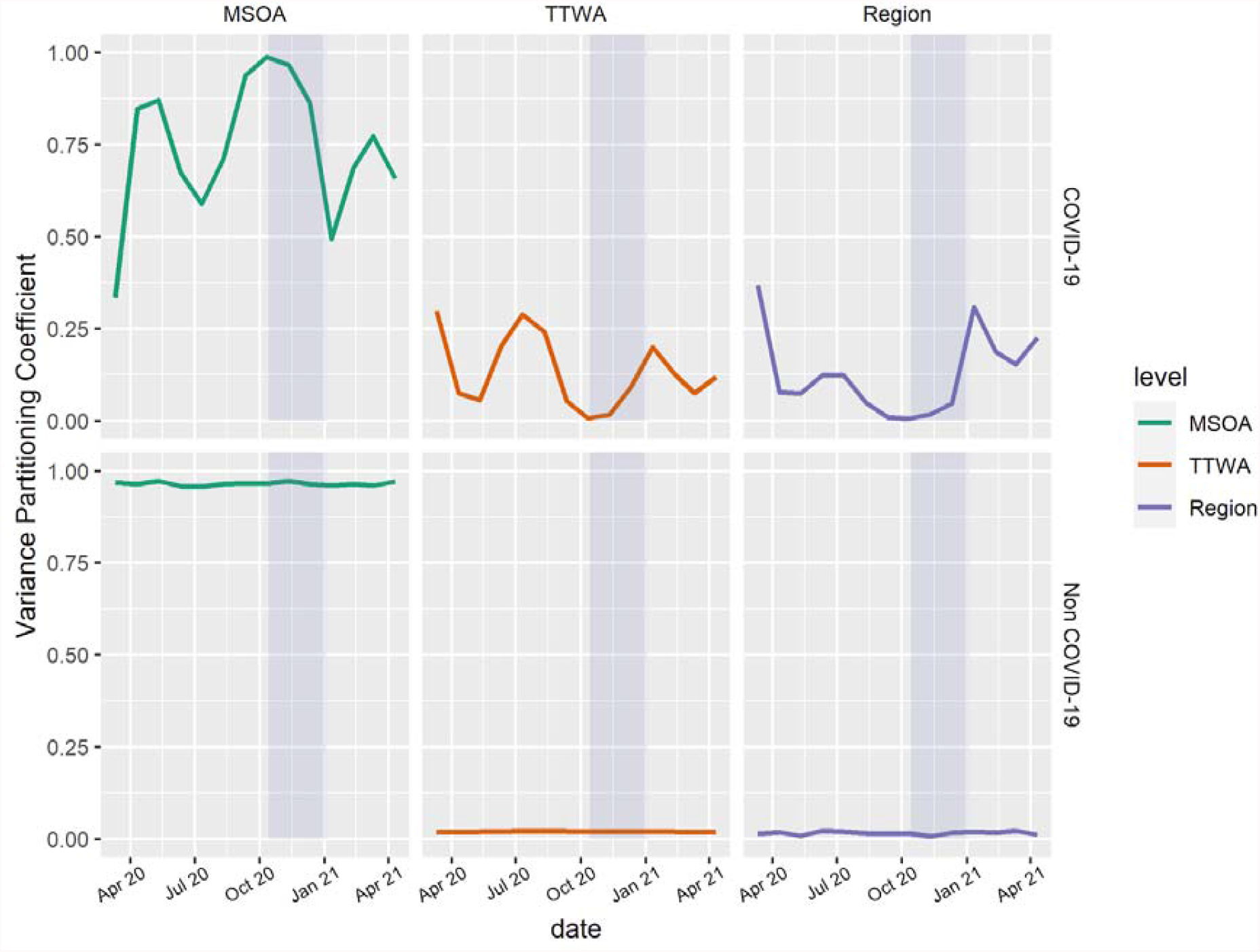
Estimates of monthly COVID-19 and non-COVID-19 VPCs across three administrative scales from March 2020 to April 2021 after adjustment for UKIMD. Purple highlight indicates the period of locally defined tiered restrictions. MSOA, Middle-Layer Super Output Area; TTWA, Travel to Work Area.

**Supplementary Figure 2.**
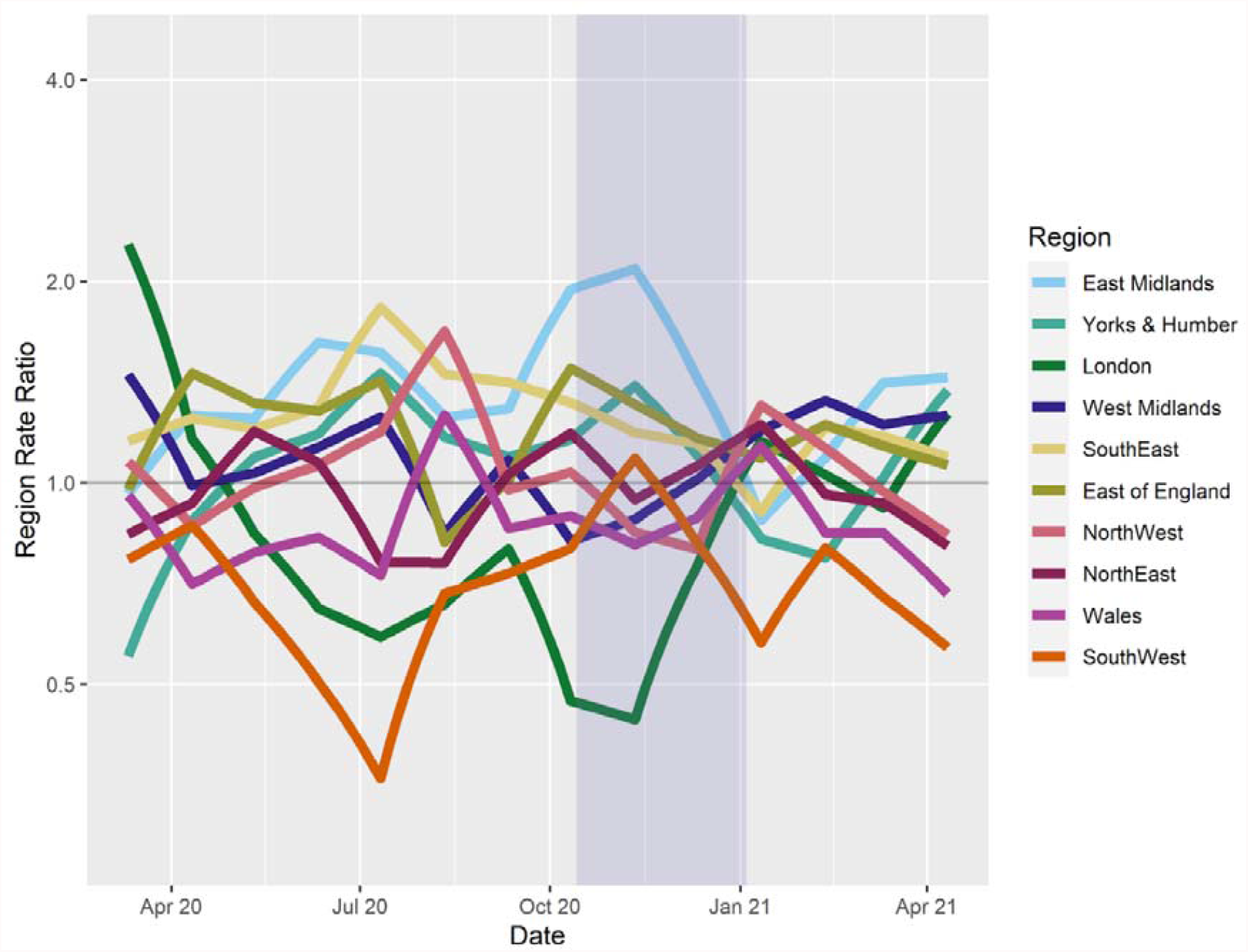
Monthly, regional COVID-19 mortality rate ratios, indicating regional mortality rate relative to precision weighted monthly population average. Model adjusted for local age structure, number of care homes and local deprivation. Purple highlight indicates the period of locally defined tiered restrictions.

## Author Contributions

GJG and GO conceived and designed the analysis and collected the data. GDS and DM provided critical feedback on the analysis design. GJG performed the statistical analysis. GJG and GO wrote the initial draft of the paper. GDS, LDH and DM provided critical revisions on the drafted manuscript.

## Acknowledgements

The Medical Research Council (MRC) and the University of Bristol support the MRC Integrative Epidemiology Unit [MC_UU_00011/1, MC_UU_00011/3].

## Data Availability

ONS Deaths due to COVID-19 mortality data are available at:

https://www.ons.gov.uk/peoplepopulationandcommunity/birthsdeathsandmarriages/deaths/datasets/deathsduetocovid19bylocalareaanddeprivation

ONS Deaths involving COVID-19 mortality data are available at:

https://www.ons.gov.uk/peoplepopulationandcommunity/birthsdeathsandmarriages/deaths/datasets/deathsinvolvingcovid19bylocalareaanddeprivation

ONS Administrative geographical identifiers are available at:

https://geoportal.statistics.gov.uk

ONS population health estimates are available at:

https://www.ons.gov.uk/peoplepopulationandcommunity/populationandmigration/populationestimates/datasets/middlesuperoutputareamidyearpopulationestimates.

ESRI care home estimates are available at:

https://covid19.esriuk.com/datasets/e4ffa672880a4facaab717dea3cdc404_0

Equivalised UK IMD estimates are available at:

https://github.com/mysociety/composite_uk_imd

All code to reproduce analyses is available at:

www.github.com/zimbabwelsh/covid_mort_spatseg

